# Basic reproduction number of 2019 Novel Coronavirus Disease in Major Endemic Areas of China: A latent profile analysis

**DOI:** 10.1101/2020.04.13.20060228

**Authors:** Honglv Xu, Min Yuan, Liya Ma, Meng Liu, Yi Zhang, Wenwen Liu, Hong Gan, Fangbiao Tao

## Abstract

**Objective:** The aim of the study is to analyze the latent class of basic reproduction number (R_0_) trend of 2019 novel coronavirus disease (COVID-19) in major endemic areas of China.

**Methods:** The provinces that reported more than 500 cases of COVID-19 till February 18, 2020 were selected as the major endemic area. The Verhulst model was used to fit the growth rate of cumulative confirmed cases. The R_0_ of COVID-19 was calculated using the parameters of severe acute respiratory syndrome (SARS) and COVID-19, respectively. The latent class of R_0_ was analyzed using a latent profile analysis model.

**Results:** The median R_0_ calculated from SARS and COVID-19 parameters were 1.84 - 3.18 and 1.74 - 2.91, respectively. The R_0_ calculated from the SARS parameters was greater than that of calculated from the COVID-19 parameters (*Z* = −4.782 - −4.623, *P* < 0.01). Both R_0_ can be divided into three latent classes. The initial value of R_0_ in class 1 (Shandong Province, Sichuan Province and Chongqing Municipality) was relatively low and decreases slowly. The initial value of R_0_ in class 2 (Anhui Province, Hunan Province, Jiangxi Province, Henan Province, Zhejiang Province, Guangdong Province and Jiangsu Province) was relatively high and decreases rapidly. Moreover, the initial value of R_0_ of class 3 (Hubei Province) was between that of class 1 and class 2, but the higher level of R_0_ lasts longer and decreases slowly.

**Conclusion:** The results indicated that overall trend of R_0_ has been falling with the strengthening of China’s comprehensive prevention and control measures for COVID-19, however, presents regional differences.

## 1. Introduction

Of particular concern is that 2019 novel coronavirus disease (COVID-19) outbroke in Wuhan, Hubei Province at the end of 2019, and quickly spread to the whole country of China (Liu et al., 2020; Lu et al., 2020). COVID-19, an infectious disease caused by 2019-nCoV, can be transmitted through droplets, aerosols, and contact (Chen et al., 2020; Li et al., 2020). The main clinical features of the case are fever, dry cough, and fatigue. There are also mild cases and asymptomatic pathogen carriers (Guo et al., 2020; Huang et al., 2020). Evidence demonstrates that COVID-19 is extremely infectious, even during the incubation period, and the entire population is susceptible (Nishiura et al., 2020; Thompson, 2020). It has a higher case fatality rate, posed great threats to public health and attracted an enormous concern (Yang et al., 2020). Currently, COVID-19 has become widespread around the world (Sommer et al., 2020; Sorbello et al., 2020). For a new infectious disease, it is important to identify the epidemic characteristics and transmission dynamics for the prevention and control of the infectious disease (Kannan et al., 2020).

It is generally known that basic reproduction number (R_0_) is an important parameter for studying the dynamics of infectious disease transmission for describing the ability to spread of an infectious source (Ganyani et al., 2018; Sato, 2019). R_0_ is defined as the average number of secondary cases that an infected subject produces over its infectious period in a susceptible and uninfected population (Chang, 2017). There has been increasing concern that R_0_ of COVID-19 is used to assess the spread of infectious diseases, predict epidemic trends, and evaluate the effectiveness of prevention and control measures (Chowell et al., 2007; Park et al., 2020). Although some previous studies have investigated the R_0_ of COVID-19, these studies are prediction estimates based on data from the early stages of the COVID-19 epidemic. Research data is very limited and results are inconsistent (Wu et al., 2020; Zhao et al., 2020; Zhao and Chen, 2020; Zhou et al., 2020). The present study analyzed the latent class of R_0_ of COVID-19 in major endemic areas that reported more than 500 cases of COVID-19 in China. We evaluate the effect of China’s prevention and control measures based on the trend of R_0_, and provide a basis for further prevention and control of COVID-19 and other emerging infectious diseases.

## 2. Materials and methods

### 2.1 Materials

The major endemic areas of COVID-19 is defined as the area where the cumulative number of cases more than 500 of COVID-19 has been diagnosed by 24:00 on February 18, 2020. These areas include Guangdong Province, Jiangxi Province, Hunan Province, Chongqing Municipality, Sichuan Province, Hubei Province, Anhui Province, Zhejiang Province, Jiangsu Province, Henan Province and Shandong Province. We collected the cumulative number of cases of COVID-19 from the official websites of the health commission in these areas. In addition, we collected the main prevention and control measures for COVID-19 from the websites of provincial governments.

### 2.2 Methods

The Verhulst model was used to fit the growth rate of cumulative number of cases (Tang et al., 2020; Zeng et al., 2003). Verhulst curvilinear equation was as follows:

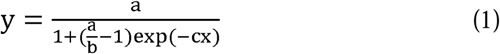

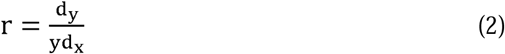

where a, b, and c are parameter constants, and r is the growth rate. We calculated the R_0_ of COVID-19 using the following equation (Wallinga and Lipsitch, 2007):

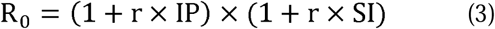

where r is the growth rate, IT is incubation period (IP), SI is the serial interval (SI) from the onset of case i to infection of case ii. We calculated R_0_ based on the IP and SI of COVID-19 (reported by the Chinese Center for Disease Control and Prevention) and severe acute respiratory syndrome (SARS). The IP and SI of COVID-19 are 5.2 and 7.5, respectively (Li et al., 2008). The IP and SI of SARS are 6.0 and 8.4, respectively (Lipsitch et al., 2003). We described the changing trend of R_0_ with time in each province, and analyzed the latent class according to the trend of R_0_. Moreover, the major prevention and control measures for COVID-19 in each major endemic area are sorted out. We used the WPS 2019 to draw a Nightingale rose diagram based on the frequency of prevention and control measures were adopted.

### 2.3 Statistical analyses

All statistical analyses were performed using R (R-3.5.1, R Core Team) and Mplus (Mplus version 7.4). Methods for analyzing include descriptive statistical analysis and model estimation. The Verhulst model was used to fit the growth rate of the cumulative number of cases and calculate R_0_ in R software. Use the range between the minimum and maximum values, the 25th percentile (*P*_*25*_), the 50th percentile (*P*_*50*_), and the 75th percentile (*P*_*75*_) to describe the distribution of R_0_. The Wilcoxon signed-rank test was used to compare the differences of R_0_ calculated from the COVID-19 parameter and the SARS parameter. A latent profile analysis (LPA) was used to analyze the latent class of R_0_ trend in Mplus software.

## 3 Results

### 3.1 The trend of the cumulative number of cases

**Figure 1** shows the trend of the cumulative number of COVID-19. It shows S-shaped growth. Hubei Province has the largest cumulative number of cases and the fastest growth. Except Hubei Province, the trends of the cumulative number of cases in several other provinces were observed in the phenomenon of classification. The number of cases in class 1 (Henan Province, Zhejiang Province, and Guangdong Province) was relatively large and has increased rapidly. The number of cases in class 2 (Anhui Province, Hunan Province and Jiangxi Province) and the growth rate were at a medium level. The number of cases in class 3 (Shandong Province, Jiangsu Province, Sichuan Province, and Chongqing Municipality) and the growth rate were relatively low. In general, except Hubei Province, the major epidemic areas of COVID-19 in China reached the highest peaks of the “S” curve around February 15.

**Figure 1.**
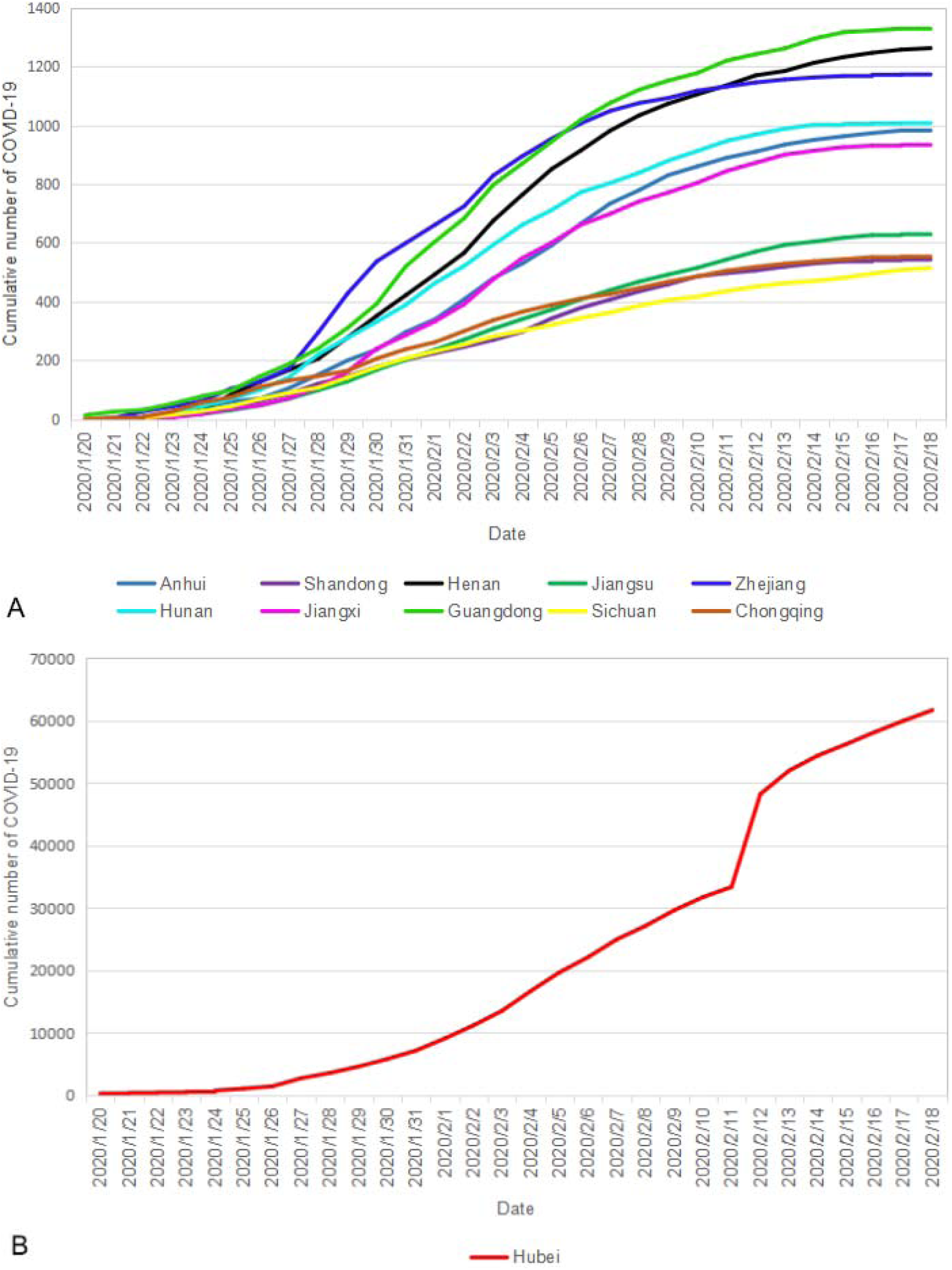
The cumulative number of COVID-19 in the major epidemic areas of China.

### 3.2 The growth rate trend of the cumulative number of cases

The growth rate of cumulative number of COVID-19 in Hubei Province is 0.03 - 0.20. Especially, the growth rate of 0.24 continued until January 26, 2020, and the growth rate of 0.23 - 0.22 continued until January 31, 2020. Then, the growth rate shows a clear downward trend in Hubei Province. Except Chongqing Municipality, Sichuan Province, and Shandong Province (the growth rates were 0.14 - 0.20), the initial growth rates (0.25 - 0.32) of all other provinces were higher than those of Hubei Province, but it started to decline rapidly after 2 - 3 days. (**Figure 2**)

**Figure 2.**
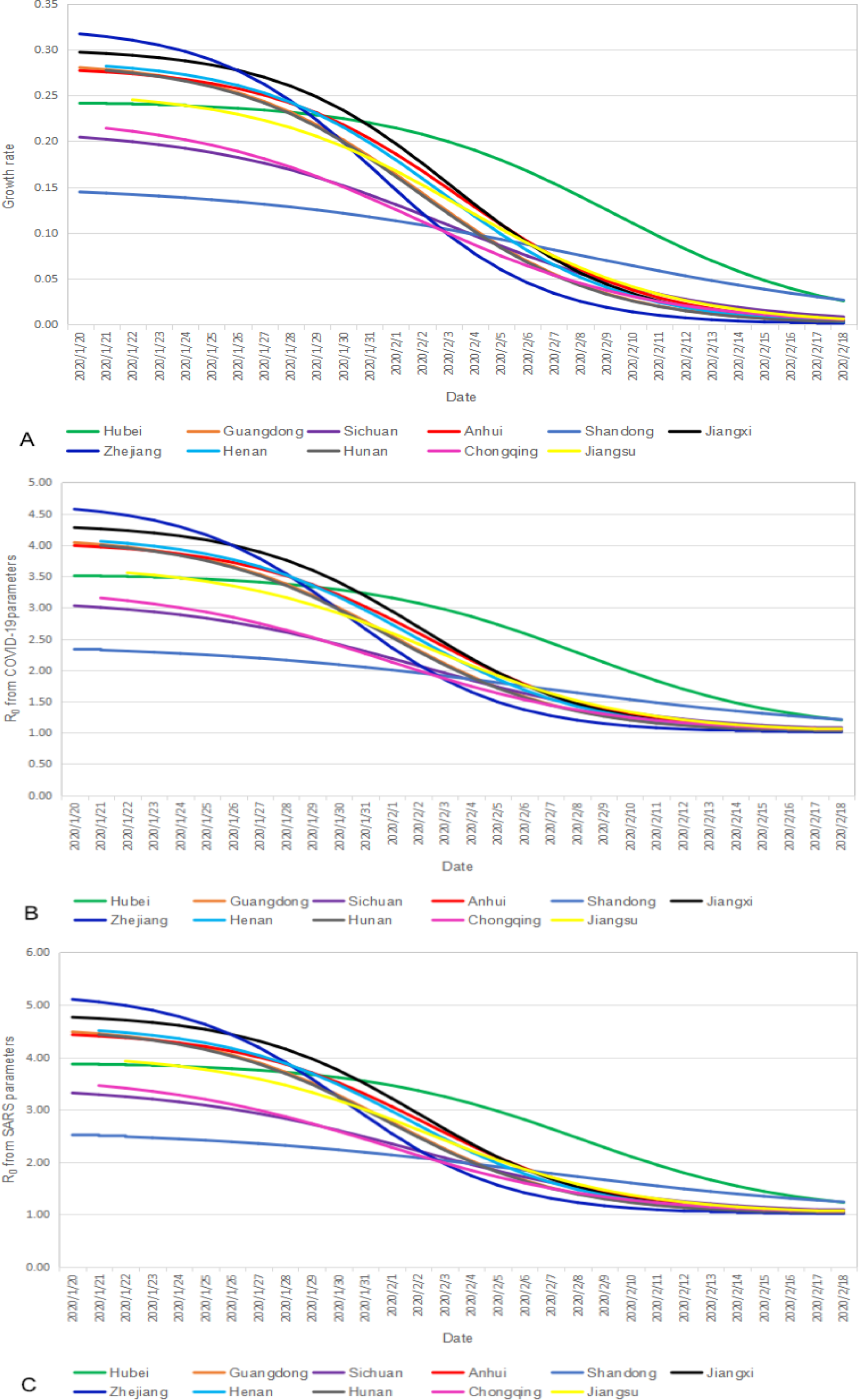
The trend of growth rate and R0.

### 3.3 The R_0_ trend of COVID-19

**Table 1** shows the distribution of R_0_. The median R_0_ calculated from SARS parameters is 1.84 - 3.18, and the R_0_ calculated from COVID-19 parameters is 1.74 - 2.91. Overall, the former is greater than the latter (*Z*=-4.782 --4.623, *P*< 0.01). **Figure 2** shows the R_0_ trend of COVID-19. The trends of R_0_ calculated from the two parameters are basically consistent. It was showed a gradual downward trend of R_0_ in each province from January 20 to February 18, 2020. The R_0_ in Hubei Province declined slowly before January 31, and then showed a significant downward trend. Except Chongqing Municipality, Sichuan Province, and Shandong Province, the initial R_0_ in all other provinces was higher than that of Hubei Province, but the decline rate of R_0_ is higher than Hubei Province. As of February 18, 2020, except Hubei Province (R_0_ = 1.20) and Shandong Province (R_0_ = 1.21), the R_0_ (1.01 - 1.06) were close to 1 in all other provinces.

**Table 1.** The distribution of R_0_ of COVID-19

| Province | $R_0$ from SARS parameters | | | | $R_0$ from COVID-19 parameters | | | | $Z$ | $P$ |
| --- | --- | --- | --- | --- | --- | --- | --- | --- | --- | --- |
| | Range | $P_{25}$ | $P_{50}$ | $P_{75}$ | Range | $P_{25}$ | $P_{50}$ | $P_{75}$ | | |
| Hubei | 1.23-3.87 | 1.90 | 3.18 | 3.76 | 1.20-3.51 | 1.80 | 2.91 | 3.41 | -4.782 | <0.01 |
| Guandong | 1.02-4.48 | 1.16 | 2.13 | 3.93 | 1.02-4.04 | 1.15 | 2.00 | 3.56 | -4.782 | <0.01 |
| Sichuan | 1.07-3.32 | 1.28 | 2.01 | 2.94 | 1.06-3.03 | 1.25 | 1.90 | 2.71 | -4.782 | <0.01 |
| Anhui | 1.04-4.43 | 1.24 | 2.44 | 4.03 | 1.03-3.99 | 1.22 | 2.26 | 3.65 | -4.782 | <0.01 |
| Shandong | 1.24-2.51 | 1.53 | 1.99 | 2.36 | 1.21-2.33 | 1.47 | 1.88 | 2.20 | -4.782 | <0.01 |
| Jiangxi | 1.03-4.76 | 1.21 | 2.49 | 4.34 | 1.03-4.28 | 1.19 | 2.31 | 3.92 | -4.782 | <0.01 |
| Zhejiang | 1.01-5.10 | 1.08 | 1.85 | 4.25 | 1.01-4.57 | 1.07 | 1.75 | 3.84 | -4.782 | <0.01 |
| Henan | 1.03-4.51 | 1.19 | 2.19 | 3.96 | 1.03-4.06 | 1.17 | 2.05 | 3.59 | -4.703 | <0.01 |
| Hunan | 1.02-4.43 | 1.15 | 2.00 | 3.78 | 1.02-4.00 | 1.13 | 1.88 | 3.43 | -4.703 | <0.01 |
| Chongqing | 1.05-3.46 | 1.20 | 1.84 | 2.93 | 1.04-3.15 | 1.18 | 1.74 | 2.69 | -4.703 | <0.01 |
| Jiangsu | 1.05-3.92 | 1.25 | 2.12 | 3.43 | 1.05-3.56 | 1.22 | 1.99 | 3.13 | -4.623 | <0.01 |

### 3.4 The latent class of R_0_

We constructed five latent class models in the latent profile analysis. The three-class model was finally selected according to the model fitting index, latent class probability, and the interpretability of the model. Table 2 shows the fitting index and latent class probability of the model. There are three latent classes of the R_0_ calculated from SARS parameters and COVID-19 parameters. Class 1 includes Shandong Province, Sichuan Province and Chongqing Municipality. Class 2 includes Anhui Province, Hunan Province, Jiangxi Province, Henan Province, Zhejiang Province, Guangdong Province and Jiangsu Province. Class 3 includes Hubei Province. **Figure 3** shows the R_0_ trend in different latent classes. The initial value of class 1 R_0_ was relatively low and decreases slowly. The initial value of class 2 R_0_ was relatively high and decreased rapidly. The initial value of class 3 R_0_ was between class 1 and class 2, but high levels of R_0_ lasted longer and declined slowly.

**Table 2.** The fitting index and latent class probability of the model

| $R_0$ | Model | AIC | BIC | aBIC | Entropy | Class probability (%) |
| --- | --- | --- | --- | --- | --- | --- |
| $R_0$ from SARS parameters | 1 | 317.51 | 341.38 | 160.72 | - | 1 |
|  | 2 | 72.44 | 108.65 | -165.35 | 1.00 | 18.2/81.8 |
|  | 3 | -92.09 | -43.55 | -410.89 | 1.00 | 27.3/63.6/9.1 |
|  | 4 | -30.09 | 30.79 | -429.90 | 1.00 | 27.3/31.8/31.8/9.1 |
|  | 5 | -83.25 | -10.04 | -564.06 | 1.00 | 18.2/27.3/36.4/9.1/9.1 |
| $R_0$ from COVID-19 parameters | 1 | 229.47 | 253.35 | 72.69 | - | 1 |
|  | 2 | -16.12 | 20.09 | -253.91 | 1.00 | 18.2/81.8 |
|  | 3 | -181.98 | -133.43 | -500.78 | 1.00 | 27.3/63.6/9.1 |
|  | 4 | -119.98 | -59.10 | -519.78 | 1.00 | 27.3/63.6/4.5/4.5 |
|  | 5 | -149.15 | -75.9 | -629.96 | 1.00 | 0/27.3/36.4/27.3/9.1 |

**Figure 3.**
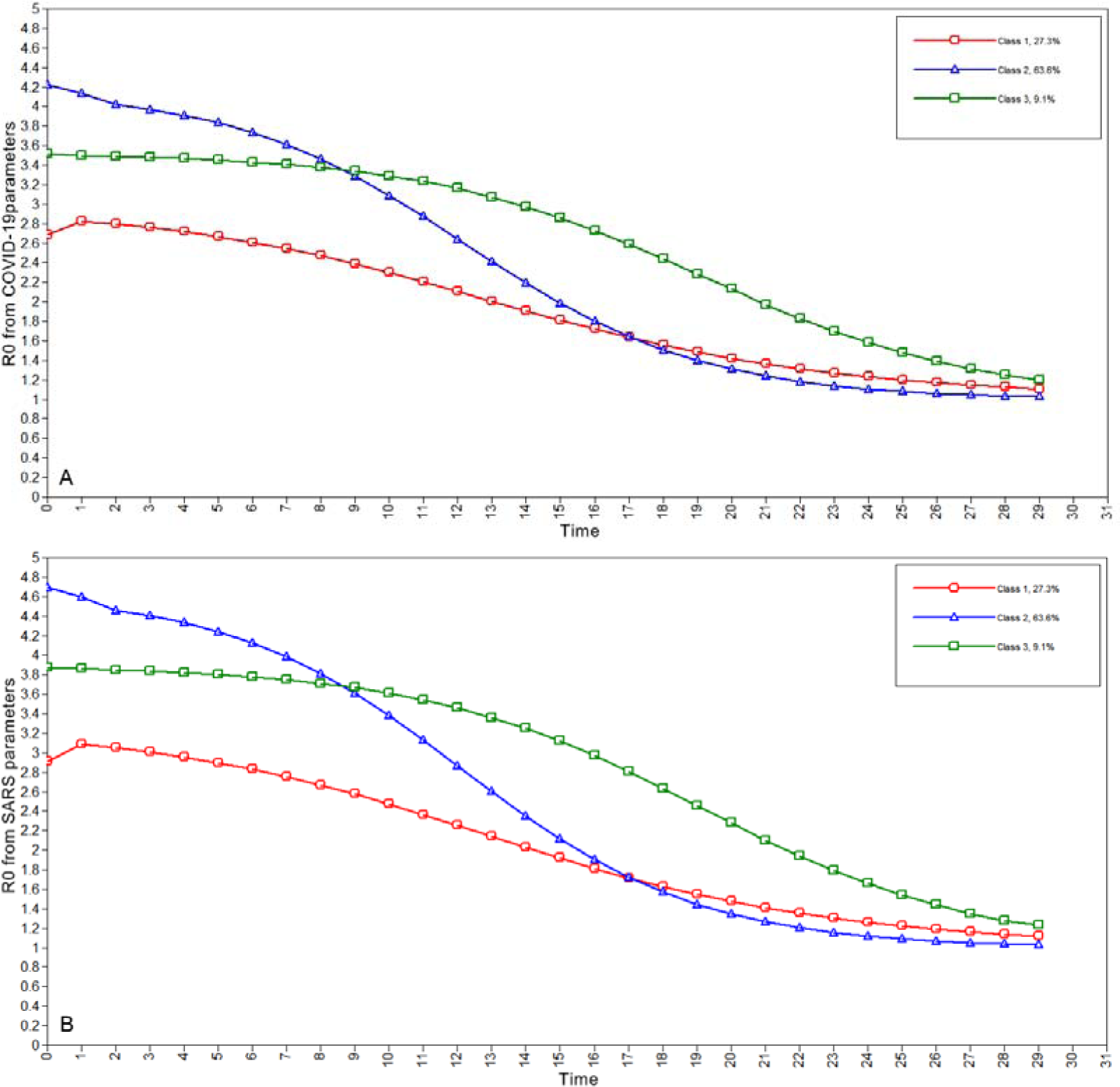
The latent class of R_0_.

### 3.5 The main prevention and control measures for COVID-19

**Figure 4** shows the main prevention and control measures were adopted for COVID-19 in major endemic areas of China. These measures include activating first-level public health emergency response, wearing masks, conducting epidemiological investigations, screening of key populations, temporary traffic control, closing public places (e.g. cinemas, internet cafes, etc.), monitoring body temperature, symptom screening, medical observation, the “four early” measures (early detection, early reporting, early isolation and early treatment), the “four concentrated” treatment measures (concentrated cases, concentrated experts, concentrated resources, concentrated treatment), quarantine oneself, lockdown a city, disinfection in public areas, and so on.

**Figure 4.**
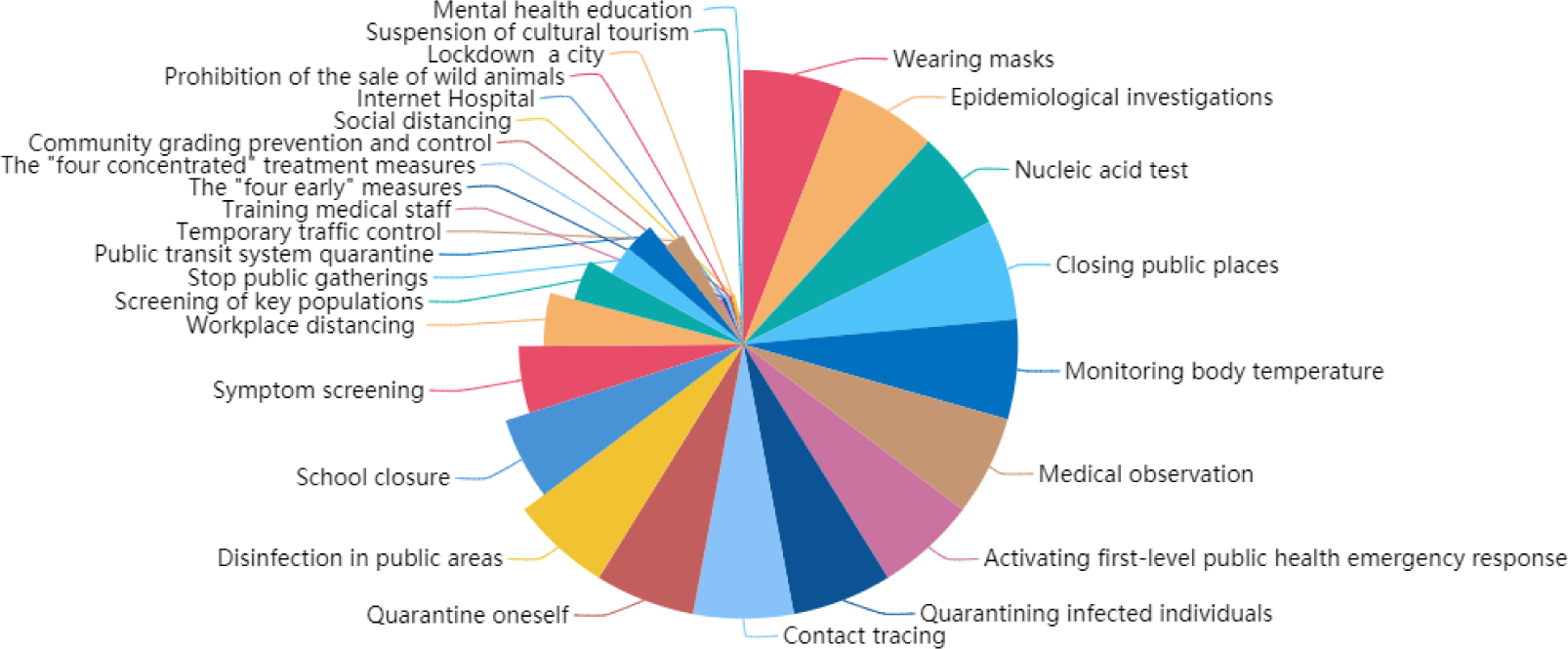
The main prevention and control measures for COVID-19 in China.

## 4. Discussion

The present study suggests that the R_0_ of COVID-19 calculated from SARS parameter is greater than the R_0_ calculated from COVID-19 parameters (the median of R_0_ is 1.84 - 3.18 and 1.74 - 2.91, respectively). Our findings may indicate that the epidemiological characteristics of COVID-19 and SARS are different. Coronavirus has caused three large-scale epidemics in humans in the past 20 years, including SARS in 2002, Middle East respiratory syndrome (MERS) in 2012 and COVID-19 in 2019 (Dye and Gay, 2003; Zaki et al., 2012). From the number of infections, deaths, and epidemic areas, COVID-19 threatens human health more than SARS and MERS (Memish et al., 2020; Sayed et al., 2020; Wilder-Smith et al., 2020). The R_0_ of COVID-19 in our study is lower than that of SARS (3.1 - 4.2) and MERS (2.0 - 6.7) (Wallinga and Teunis, 2004; Majumder et al., 2014; Chang, 2017). These findings are consistent with previous studies on the infectious capacity of COVID-19 is lower than that of SARS and MERS (Zhou, et al., 2020). Additionally, the R_0_ calculated from the COVID-19 parameters in this work is similar to the R_0_ (2. 38 - 2.72, 2.47 - 2.86) estimated in China Other studies in China (Geng et al., 2020; Wu, et al., 2020). Also, it is also similar to R_0_ (2.4 - 2.8) of COVID-19 in Japan (Kuniya, 2020). Nevertheless, our calculated R_0_ is lower than that estimated using the epidemic data in the early epidemic of the COVID-19 in Wuhan (R_0_ = 1.4 - 3.9), Hubei Province (R_0_ = 2.80 - 4.48), and China (2.8 - 3.3) (Li, et al., 2008; Wang, et al., 2020; Zhou, et al., 2020).

On the whole, R_0_ of COVID-19 in each province is gradually decreasing. It is believed that the powerful combined prevention and control measures adopted for COVID-19 by the Chinese government have had obvious effects. These combined measures and the successful experience prevention and control for COVID-19 of China can provide a reference for other countries. There is mounting evidence that implementing the combined measures could significantly reduce the number of cases such as workplace distancing and school closure, quarantining infected individuals and their family members, social distancing and quarantine measures (Anderson et al., 2020; Koo et al., 2020). A study in Singapore showed that the combined intervention will reduce the estimated number of infections by 99.3%, 93.0%, and 78.2% when R_0_ was 1.5, 2.0, and 2.5, respectively, compared with the baseline scenario (Koo, et al., 2020). Some researchers believe that isolation of cases, contact tracing, social distancing are used to control outbreaks of infectious diseases (Anderson, et al., 2020). Specifically, contact tracing is a key activity in reducing the spread of the epidemic. A new study from the UK suggests that to control the majority of COVID-19 outbreaks, for an R_0_ of 3.5 more than 90% of contacts had to be traced, and for R_0_ of 2.5 more than 70% of contacts had to be traced (Hellewell et al., 2020).

The results of the latent profile analysis showed that there are three latent class of R_0_, and the trends of R_0_ in each latent class have their own unique characteristics. Observed findings indicate that although the Chinese government has adopted the overall prevention and control measures of “a board of chess in China, suit one’s measures to local conditions”, the effects are not entirely consistent. More specifically, the initial value of R_0_ in class 1 was relatively low, and the decline was slow. The possible explanations are that these areas have taken active measures after the COVID-19 epidemic in Wuhan, and the spread of COVID-19 was well controlled from the beginning. The initial value of R_0_ in class 2 was relatively high, even higher than that of Hubei Province. However, the R_0_ declined rapidly, and the decline rate was the fastest of the three latent classes. The possible reason is that the measures adopted at the beginning of the epidemic of COVID-19 were not effective. Then, the epidemic was effectively controlled after quickly adjusting the measures in these areas, and the infectious capacity of COVID-19 decreased rapidly. The initial value of R_0_ in class 3 (Hubei Province) was between that of type 1 and type 2, but the higher level of R_0_ lasts longer and decreases slowly. Wuhan City, Hubei Province, is the first area in China where COVID-19 was found to be endemic. Due to the lack of understanding of the emerging infectious disease pathogens, transmission routes, susceptible populations, disease characteristics, epidemic characteristics, and other unknown reasons, the R_0_ of COVID-19 lasted longer at higher levels in Hubei Province. In other words, before January 31, Hubei Province COVID-19 was strongly infectivity. Therefore, our findings agree that the adoption of active prevention and control measures at the beginning of the epidemic can effectively control the spread of COVID-19.

What’s more, the R_0_ of COVID-19 in Hubei Province and Shandong Province were around 1.2 by February 18, and there is still a certain risk of transmission. The R_0_ (1.01 - 1.06) in other provinces were close to 1, and still greater than 1. It is widely recognized that R_0_ can reflect the epidemic trend of infectious diseases. When R_0_ > 1, the greater the R_0_, the greater the infectious ability of infectious diseases and the greater the number of cases. When R_0_ < 1, means the epidemic of infectious diseases will gradually end. The optimal targeting of taking effective interventions is to control the R_0_ less than 1 (Delamater et al., 2019). At present, China has gradually resumed work and school, and population movement and crowd accumulation have further increased the risk of COVID-19 transmission and the difficulty of prevention and control. Furthermore, COVID-19 has been endemic and spread in many countries around the world, and international import cases may cause a secondary epidemic (Lai et al., 2020; Palacios et al., 2020). Consequently, China also needs to continue to strengthen its prevention and control for COVID-19.

The present study had several strengths and limitations. The main strength of this work was to calculate the R_0_ based on the epidemic data of major epidemic areas in China as of February 18. However, previous research on R_0_ was almost prediction and estimation based on the limited epidemiological data of COVID-19 in Wuhan in the early days. Another strength was to analyze the latent class of R_0_ and explore the regional differences in prevention and control effects. The findings presented in this study have implications for the preliminary evaluation of the effectiveness of prevention and control measures for COVID-19 adopted in China. In addition, the research results can provide experience for other countries to response to COVID-19, and also provide support for previous prediction studies. There are several limitations to our study. Above all, the calculation of R_0_ is affected by multiple parameters and various factors (Delamater, et al., 2019). And yet the factors considered are relatively single in this study. Additional research is necessary to confirm the accuracy of the R_0,_ considering the large uncertainties around estimates of R_0_ and the duration of infectiousness (Rebuli et al., 2018; Prem et al., 2020). Additionally, there are many methods for calculating R_0_ in the world (Chowell, et al., 2007), but this study did not use multiple methods to compare the results. Finally, although this study sorts out the prevention and control measures for COVID-19 in major endemic areas in China, it is difficult to quantitative evaluate the effects of various measures.

## 5. Conclusion

Overall, we found that the R_0_ of COVID-19 shows a downward trend in major endemic areas in China, and there are regional differences (three latent classes). Actively adopting combined prevention and control measures in the early stages of the epidemic can effectively control COVID-19.

## Data Availability

The data has been shared in Mendeley Data.
http://dx.doi.org/10.17632/48j3z3fxw6.2

## Author contributions

Fangbiao TAO designed the study. Honglv XU took primary responsibility for writing the manuscript, managed the literature searches and analyses, and undertook the statistical analysis. Min Yuan, Liya Ma, MengLiu, Yi Zhang, Wenwen Liu and Hong Gan undertook the acquisition of the data. All authors contributed to and approved the final manuscript.

## Acknowledgements

This work was supported by the Anhui Medical University Emergency Key Research Project for Novel Coronavirus Pneumonia (YJGG202001) and Emergency Research Project, Anhui Provincial Department of Science and Technology, Anhui Provincial Health Commission (202004a07020002).

